# A medical algorithmic audit framework for evaluating the safety, equity, and quality of an AI Scribe tool in a paediatric developmental assessment clinic

**DOI:** 10.1101/2025.09.30.25336928

**Authors:** Melissa D McCradden, Sheng Tng, Deepa Jeyaseelan, Cathy Leane, Marnie Campbell, Timothy Braund, Lana Earle-Bandaralage, Mary Ebrahimi, Ashish Sharma, Jonathan Tang

## Abstract

**Background:** Any tool that can reduce the administrative burden on healthcare providers while preserving safe, accountable and high-quality medical documentation is of immense value both to healthcare institutions and consumers. The key question we need to answer is whether a prospective tool can reduce these burdens while maintaining (and, ideally, *elevating*) quality documentation standards.

The goal of this study is to establish whether a large language model (LLM)-based documentation assistive tool can maintain safe, high-quality documentation while reducing the time required to ensure high-quality clinical documentation in the Child Development Unit (CDU) at the Women’s and Children’s Hospital (WCH).

**Methods:** Using an algorithmic audit framework developed specific to our context, we will compare clinician-written clinical notes to AI-generated notes produced in parallel to the standard of care (i.e., a ‘silent’ or translational trial paradigm). We will compare the time required to review clinical documentation per the standard of care compared with the AI-supported workflow with consideration to the accuracy of the final documentation. Finally, we will qualitatively describe AI-generated notes and compare them to the current standard to identify specific areas where clinical guidelines (e.g., performance information, risk mitigation) would support appropriate clinical use.

**Significance:** The contributions of our protocol are twofold. First, we close a key gap in the literature which has limited attention thus far to measuring the real-world clinical utility of AI Scribes. Second, we adapt an established framework for medical algorithmic auditing to the specific use case of LLM-enabled AI Scribes. Our multi-disciplinary research team comprising clinicians, consumers, Aboriginal Health representatives, and an ethicist is unique within this research field and will allow us to generate insights from these diverse perspectives that together can provide constructive guidance for clinicians using and testing AI Scribe tools.

## INTRODUCTION

There is immense interest in the potential for generative artificial intelligence (GenAI) such as large language models (LLMs) for alleviating healthcare-related documentation burdens [1-4]. AI Scribes in particular have shown promise as tools which can reduce the time to complete clinical notes, provide discharge summaries and patient letters, or writing letters to schools, insurance companies, and other institutions [5].

AI Scribes fall into a challenging territory of evidentiary and regulatory foundations. While administrative functions in healthcare are not often considered ‘interventions’ as they are typically used to improve a current standard, it is very clear at this point that AI Scribes may introduce unique and unanticipated risks into clinical environments [6-10]. As devices, some AI Scribes use standardized prompts which the user cannot modify, thus reducing their ability to ‘provide medical advice’ - the general line that must be crossed to fall under the regulatory scope[11]. Nonetheless, ethically, it is incumbent upon the healthcare profession to use more than ‘likeability’ as a metric of whether the use of such a resource-intensive tool is justified [12]. But what kind of evidence should we use to inform decisions about implementation?

It has been highlighted that as rapidly as AI Scribe adaptation is occurring, there is still a lack of sociotechnically-informed evaluative frameworks[5]. Much of the focus has been on factual fidelity in relation to the original transcript and general quality metrics (e.g., fluency, completeness)[6]. These metrics are important, but they focus on the evaluation of medical documents as textual corpora; medical documentation is a sociotechnical phenomena, situated within a larger context of accountabilities, communicative intentions, and purposes. To assess the suitability of AI Scribes in context, we must align their evaluation within the context of their wider importance.

We propose that the methodology of the medical algorithmic audit [13] provides relevant, useful, context-specific information to inform the decision about procurement, use, and monitoring of AI Scribes. Our team at the Women’s and Children’s Health Network’s Child Development Unit - which includes physicians, consumers, Aboriginal Health representatives, and an ethicist - co-developed an evaluative approach aligned to the algorithmic audit framework. This paper describes our protocol for testing i-scribe, an Australian-developed AI Scribe device, for its functionality related to generating clinical notes for child and adolescent patients presenting for an initial assessment of neurodevelopmental concerns. We will use this methodology to test the tool ‘silently’ - here, meaning that the AI-generated notes are sent to the research team and not used for the documentation of current patient cases. This silent (or, translational) trial methodology enables the real-life local testing of an AI Scribe tool in its intended use setting without yet introducing the potential risk to patients/participants[14].

## METHODS

### Patient and Public Involvement

In preparation for this project, the team includes four consumer partners (AS, LE, ME, TB) under the ‘collaborate’ model for patient engagement in health research [15]. Under this model, these individuals are members of the research team and have taken an active role in shaping this protocol, the evaluative framework, and will take part in the data analysis and interpretation of the research outcomes.

### Consultation and Partnership with Aboriginal Health

Relevant within our context is a commitment to equity and Aboriginal rights. While some report on diagnostic biases of LLMs, no studies have yet addressed the cultural appropriateness and suitability with respect to generating text related to clinical documentation for First Nations peoples.

In South Australia (SA), a key guiding document is the SA Health Aboriginal Impact Policy [16] in addition to our network’s/ hospital’s Aboriginal Health Plan [17]. Under SA Health Policy, when a research proposal is linked with Aboriginal-specific initiatives and has a high impact to Aboriginal peoples, consultation is required to proceed. Our research team reflected that based on the literature on LLMs in healthcare, major risks to bias have been identified specific to marginalised groups [18-21]. We recognised that there would be a need to evaluate the AI Scribe specifically on its quality and safety with respect to Aboriginal consumers and therefore needed to consult the Aboriginal Health team at WCHN.

Building on an ongoing relationship between the Aboriginal Health team and the AI Director at WCHN, we have explored the potential benefit of ‘generic’ text for describing clinical visits and how this might reduce problematic descriptions of Aboriginal consumers in clinical notes [ref]. At the same time, the group also recognises that technologies trained on dominant data sources will not likely be able to reproduce culturally appropriate descriptions and may not capture the cultural ways of conversing in a correct manner, and so there may be more risks of incorrect documentation for Aboriginal consumers. We cannot be sure until we test these AI tools, and so the focus must be on minimising risk to consumers and integrating cultural safety and respect in this protocol. To follow a culturally respectful engagement protocol, the non-Aboriginal team members followed the guidance of the South Australian Health and Medical Research Institute(SAHMRI) Research Accord [22] which provides a set of principles for culturally responsive research partnerships. See Table 1 for a representation of how we operationalised these principles in the CDU AI Scribe Project.

**Table 1:** Instituting SAHMRI’s Research Accord Principles.

| Table 1: Instituting SAHMRI’s Research Accord Principles |
| --- |
| **Priorities:** The study addresses a recognised need to improve clinical documentation for Aboriginal consumers. The Aboriginal Health and Wellbeing Division reports ongoing challenges with respect to clinical documentation in healthcare contexts. There is alignment between this project and the Aboriginal Health Plan (2024-2028).<br>**Involvement:** The Aboriginal team members are actively engaged both in this project and broader AI strategy development, including its conceptualisation, design, conduct, and interpretation. These team members will lead the evaluation with respect to bias and will have full authority over the communication of any findings relating to the implications for Aboriginal consumers.<br>**Partnership:** This specific research project builds on an existing relationship between the Aboriginal Health and Wellbeing Division at WCHN and the AI team involving ongoing partnership around all matters relating to AI at the institution and across the network.<br>**Respect and Communication:** All non-Aboriginal team members have undertaken cultural training and defer to Aboriginal CIs on culturally specific matters. Non-Aboriginal team members maintain a commitment to receiving feedback to maintain a culturally inclusive research environment.<br>**Reciprocity and Ownership:** Findings will be used to reduce bias in AI-generated clinical documentation, with benefit-sharing and protection of intellectual property ensured. Decisions about authorship will be made collaboratively and honour the work and roles of team members.<br>**Control:** Aboriginal investigators will lead the interpretation of data relating to Aboriginal |

### Participants, Setting, and Recruitment

Upon initiation of the study, all patients (consecutive recruitment) and families who are being scheduled for an initial Paediatric Assessment will be approached about participation. A

Paediatric assessment in the CDU typically involves a child who is being assessed for neurodevelopmental conditions and includes a developmental assessment and physical examination. Given that the protocol involves transcription of all verbal utterances within the encounter, each individual present in the visit will be approached for consent/assent. We will screen using the following inclusion criteria: (1) visit purpose is for an initial Paediatric assessment; (2) able to conduct the visit in English; (3) the patient is between 0-16 years of age;(4) all persons present for the visit provide consent or assent to the study. Exclusion criteria will be applied as follows: (1) families where the primary historian requires an interpreter for their assessment; (2) families where there are potential safety issues or custody disputes; (3) cases where the legal guardian is not the person providing the medical history during the consult such as young people under guardianship; (4) when any individual who wishes to be present for the visit does not agree to participate in the study.

Given the use of this novel evaluative framework, on consultation with a statistician, we are not conducting an a priori sample size but instead focus on an estimated trial length of 6 weeks. The team will review the first week’s set of AI-generated notes and complete the evaluation to develop a more empirically-informed estimate of the required sample size needed for this study. We will review on an ongoing basis at an approximate period of 10 cases to ensure we can conduct secondary analyses (e.g., around fairness issues). Based on an estimate of 10-11 cases per week at minimum, we anticipate 40-44 cases per month to be eligible for the study. Previous anecdotal evidence suggests that the anticipated consent rate will be quite high (90%+), so we expect that in 2-3 months we would have close to 100 cases which should allow us to have a more than suitable sample.

At the time of booking the assessment, WCH CDU Clinical Coordinators will phone families to confirm the assessment time. During this phone call, WCH CDU Clinical Coordinators will ask if they would agree to receive a phone call from a member of the research team. A research team member will phone consenting families to explain the study, provide information, and answer questions, emphasising that participation is voluntary and that, should the family decline participation, their child’s care and relationship with the health service will not be compromised. Families will receive the study information via mail or email as per their specified preference.

Aboriginal and Torres Strait Islander consumers will be offered the opportunity to speak with an Aboriginal research team member at the first point of contact to ask any questions they have or receive clarification. As many may prefer in-person discussion about participating in the study, when this is desired the study team will arrange an in-person presence with sufficient time given prior to the visit to support the family’s choice to participate in the project or not. Potential participants were not required to attend any additional visits and there was no change to the typical care they receive for clinical care. Contact information for the study team, the Aboriginal research team members, and the Research Ethics Committee are provided.

Informed consent and assent will be obtained prior to the date of the clinic appointment, and then confirmed again on the day of the appointment. The family will be shown a short demonstration of the i-scribe AI technology at the beginning of the consultation. Once consent/assent of all parties has been confirmed by the assessing physician at the visit, the physician will collect the completed consent form. The AI tool will be activated in front of the client and their caregiver for the duration of the assessment.

### Study Procedures

The AI Scribe tool being tested is i-scribe (Akuru). i-scribe is a generative AI tool enabled through a web application which is accessible upon securing a license to the software. i-scribe functionality includes a dictaphone mode, consult recording mode, and clinical note mode to suit individual preferences on how clinicians and consumers wish to engage with the tool. i-scribe does not permit ‘text expanders’ - that is, the user’s ability to prompt the system to generate more text. The Akuru team worked with the CDU clinicians to create a custom template specific to their consultation process. The CDU team received on-site training on the product prior to the study period.

The clinician will use i-scribe to generate a Visit Report and Patient Letter and store both in a dedicated research folder under the participant’s study ID number. The AI generated transcription and report will not be reviewed by the treating clinician at the time of note generation, and they will not be included in the participant’s medical record. The study team will then de-identify the transcription, report, and letter and update the files to ensure the de-identified version is used for the study. A copy of the clinician generated report will be de-identified and included in the research study folder to enable comparison. The AI generated report will be primarily compared against the clinician generated report. Only experienced paediatricians will participate in this study as clinicians to maintain consistency of content captured.

### Evaluative framework - algorithmic audit methodology applied to an AI Scribe

The evaluative framework follows the guidelines for conducting a medical algorithmic audit which is based on the Failure Modes and Effects Analysis (FMEA) methodology[13]. The stages include: scoping, mapping, artifact collection, testing, and reflection.

#### Scoping

The audit scope includes looking at established clinical documentation standards and relevant principles that apply (Table 2). We used these documents to identify the following requirements for our evaluation: 1) fidelity of the captured information; 2) objective, person-centred language to describe the patient, family, and their behaviours; 3) both medical and social information as clinically relevant; 4) information sufficient to assist with care coordination.

**Table 2:**
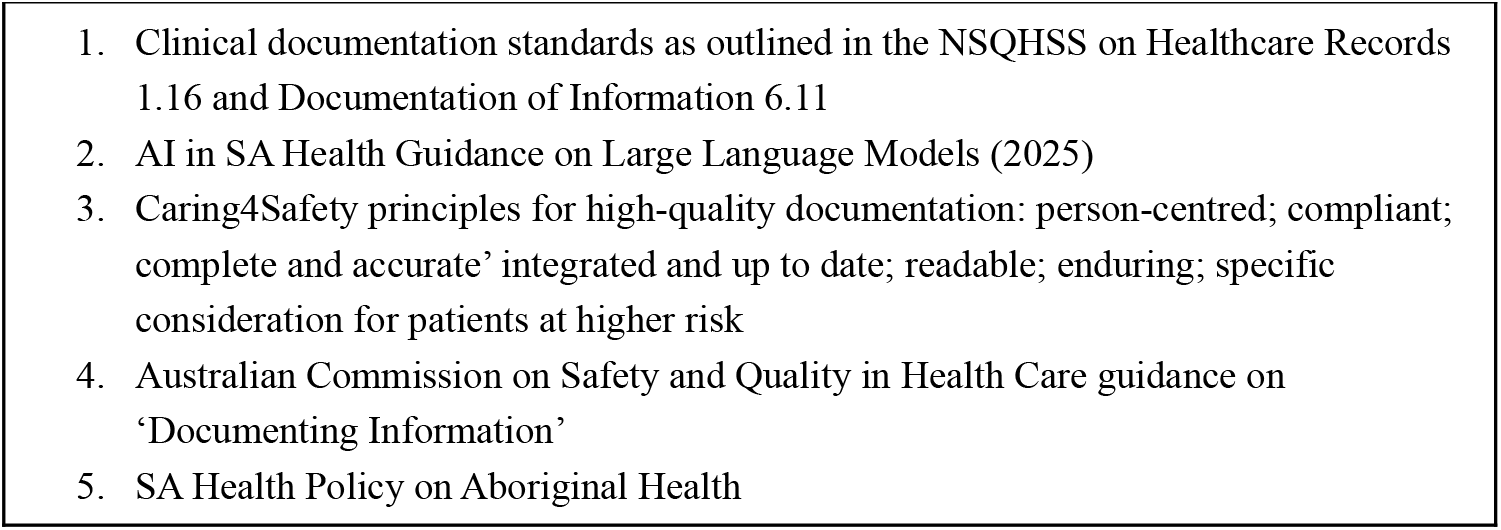
Source documents to ground out audit framework.

| Table 2: Source documents to ground out audit framework |
| --- |
| 1. Clinical documentation standards as outlined in the NSQHSS on Healthcare Records 1.16 and Documentation of Information 6.11<br>2. AI in SA Health Guidance on Large Language Models (2025)<br>3. Caring4Safety principles for high-quality documentation: person-centred; compliant; complete and accurate' integrated and up to date; readable; enduring; specific consideration for patients at higher risk<br>4. Australian Commission on Safety and Quality in Health Care guidance on 'Documenting Information'<br>5. SA Health Policy on Aboriginal Health |

#### Mapping and Artifact collection

The intended use also defines the audit scope, then mapped to risks. While traditionally a regulatory term, the intended use statement is a reliable means of defining precisely the scope for how a given product is supposed to be used. Based on the intended use, we draw from the FMEA risk priority assessment for determining risks. This includes clinically significant risks rated from 1 (minor, no intervention needed and injury to patient) to 4 (catastrophic, cause injury or death, or extreme loss of trust), and by severity, where 4 is frequent and severe and 1 is remote and benign. We also consider the ‘detection’ ability (i.e., the ability of a clinician to detect the error reliably before it results in adverse consequences) qualitatively with the clinical evaluators.

Consumers assisted the research team in determining the severity and consequences of these risks (Table 3).

**Table 3:**
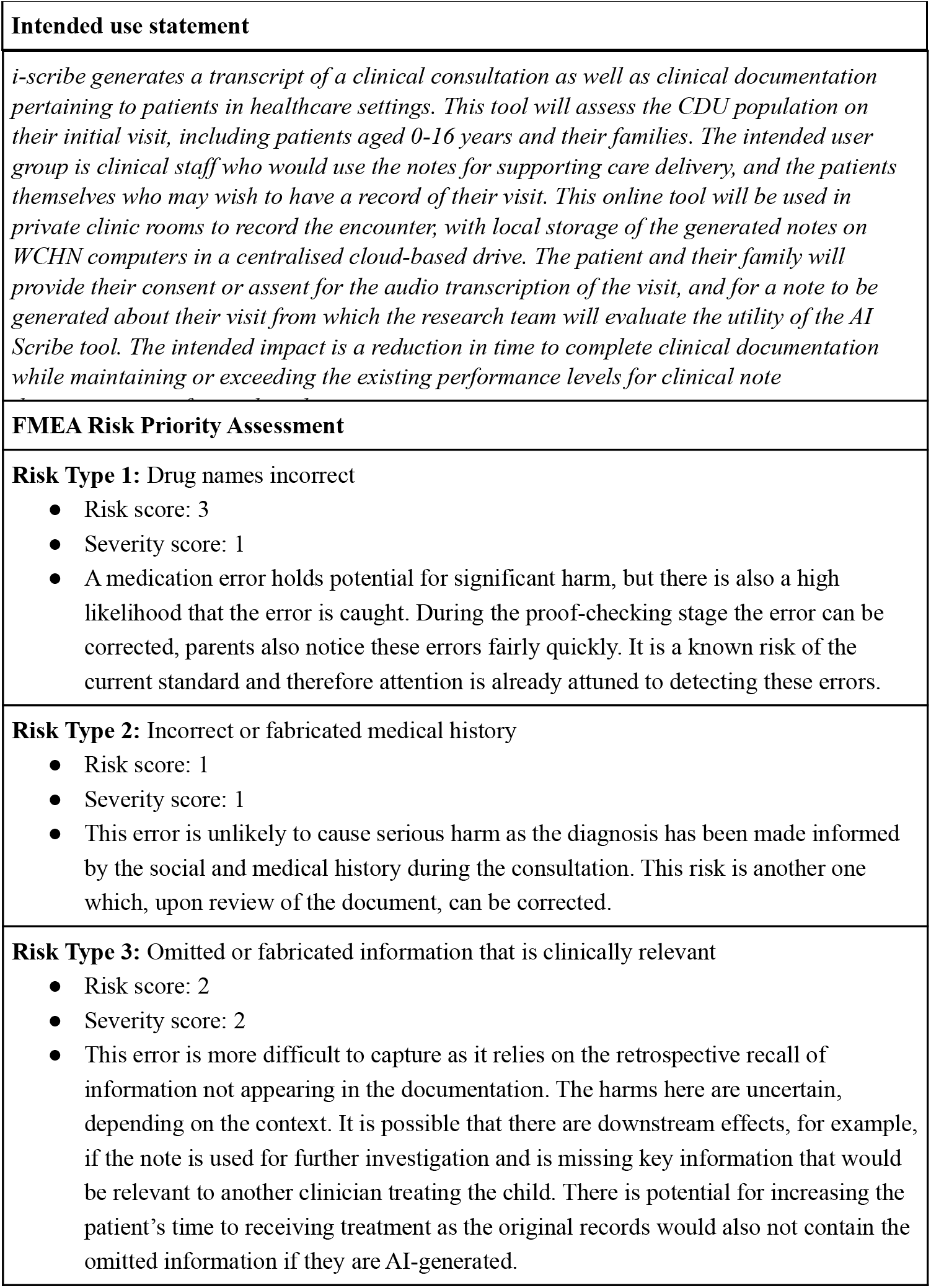
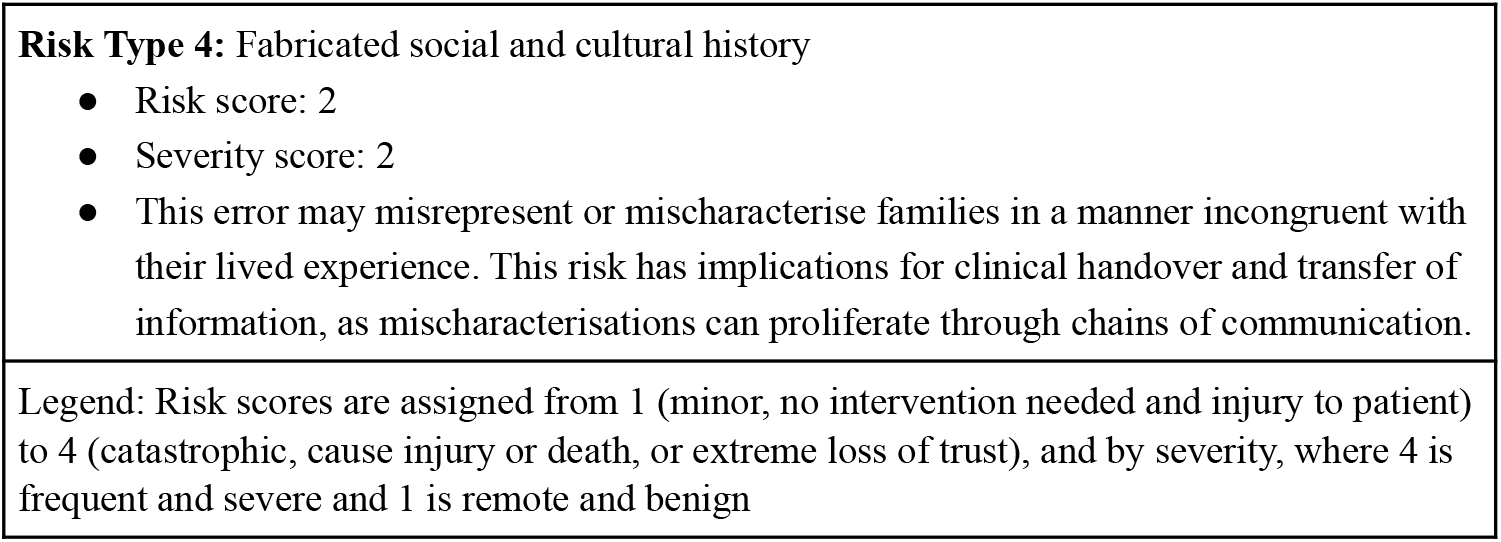
Intended use and FMEA.

#### Testing

We will conduct exploratory error analysis to describe the errors both qualitatively and quantitatively. Categorical assessment will be used for each Risk Type (e.g., ‘absent’ ‘present’ for each Risk Type) plus qualitative analysis of the nature, likelihood of detection, and clinical implications of the Risk.

To identify specific categories of risk pertinent to Aboriginal and Torres Strait Islanders consumers, we will use a deductive-inductive approach to determine whether (a) there is a significant difference in the error rate for ATSI consumers on any of the Risks above; (b) whether we find evidence of ATSI-specific biases in the generated notes.

#### ATSI-Specific Bias Assessment

The taxonomy of bias developed by Pfohl and colleagues provides a useful starting point for examining biases. Though not developed specifically for and with Aboriginal perspectives (i.e., First Nations persons residing in present-day Australia), we will capture any forms of structural bias using their framework (deductive) and emergent observations identified by the Aboriginal members of the research team (inductive) [Table 4].

**Table 4:**
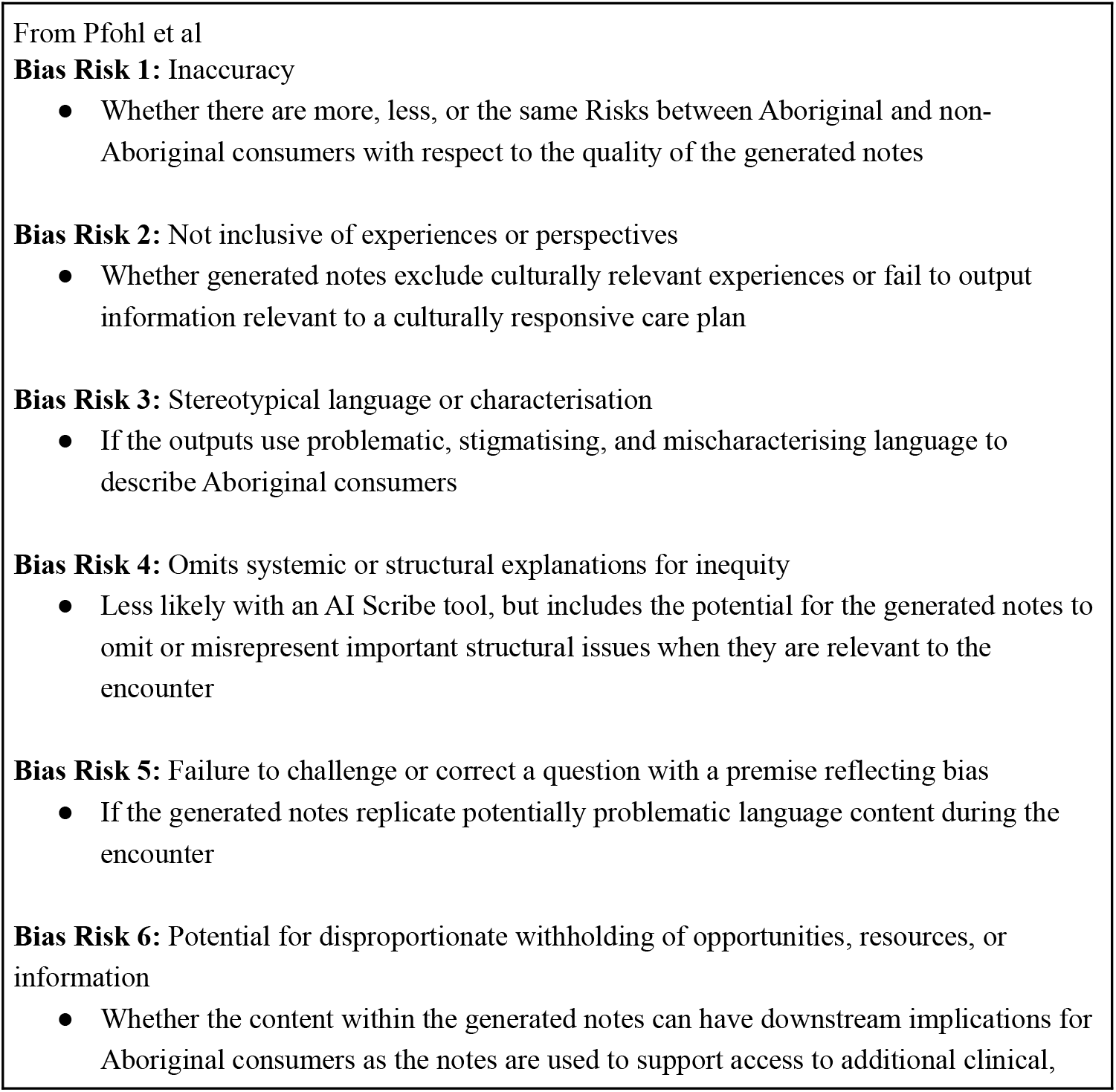
ATSI-Specific Bias Assessment.

#### Reflection and post audit processes

On the completion of the empirical evaluation of the audit, we complete the Reflection and Post Audit Processes as per the FMEA framework. This includes an assessment of the risks, risk mitigation measures, and specifying developer and clinical actions. We will undertake this reflection as a collaborative endeavour amongst our research team, and seek broader input as appropriate.

The post-audit process includes looking at developing support for clinician use of the tool, should the trial move forward (into clinical use). The results of the audit will be used to develop a framework for clinical use of i-scribe, with the goal of providing context-specific risk mitigation measures. We additionally consider the value of this work as an opportunity to enhance person-centered, culturally inclusive documentation practices to address WCHN’s Strategic Priorities of ‘Making meaningful gains in Aboriginal Health.’’ [17].

### Ethics Statement

This study has been approved by the Women’s and Children’s Human Research Ethics Committee (HREC) #00067 and the Aboriginal Human Research Ethics Committee (AHREC) #04-25-1185.

There are no anticipated benefits from this study to the participants because they are receiving care-as-usual to maintain the ‘silent’ nature of this study. A potential secondary effect is that patients may appreciate the conscientious approach to safety and quality regarding AI that is being undertaken by the team, which hopefully incurs their trust. We intend and hope to promote cultural safety in this study by attending to the Research Accord Principles to support Aboriginal and Torres Strait Islander consumers in considering participation in this study.

## CONCLUSION

As we pursue the benefits of assistive documentation using AI, we must appropriately calibrate our use of these tools according to local, context-specific information about performance. Additionally, we don’t just want automation for automation’s sake - we should be aiming for *improvement*.

Certainly, generating a clinical note at the press of a button which would otherwise have taken time for a clinician to manually write is not in question; the crucial question is whether the AI tool can do so safely, of sufficient quality, and whether the time it takes for a physician to appropriately review this note is less than the time it would take them to use a manual process or available alternatives (e.g., dictation). Taking a risk-based approach allows us to develop an empirically-grounded set of facts about the tool’s performance in its local context in order to provide constructive guidance for clinicians to address these risks while maintaining the benefits of the efficiency gains.

We hope that this protocol strengthens the discussion in the literature around the evidence-based evaluation of AI Scribe tools, and offers a positive example of a collaborative, inclusive research process that addresses the specific risks to minoritised consumers.

## Data Availability

Protocol paper, NA

https://osf.io/56cxm/

## AUTHORS CONTRIBUTIONS

All authors participated in the planning of the study, protocol development, and contributed to the development of the auditing framework. All authors have reviewed and approved the final manuscript.

## ACKNOWLEDGEMENTS

We are grateful to the support from Dianna Smith-McCue, Director of Consumer Experience at Women’s and Children’s Health Network for her assistance with connecting us with the consumers who collaborated for this project. Thank you to Dr. Emily Powell (*i-scribe*) whose support has been invaluable for getting this project off the ground. MDM wishes to thank Mr. Lukas Korkuti, a youth patient and research partner whose insights

## REFERENCES

1. Bedi S, Liu Y, Orr-Ewing L, Dash D, Koyejo S, Callahan A, Fries JA, Wornow M, Swaminathan A, Lehmann LS, Hong HJ. Testing and evaluation of health care applications of large language models: a systematic review. JAMA 2025; 333(4):319–328. doi: 10.1001/jama.2024.21700.

2. Van Veen D, Van Uden C, Blankemeier L, Delbrouck JB, Aali A, Bluethgen C, Pareek A, Polacin M, Reis EP, Seehofnerová A, Rohatgi N. Adapted large language models can outperform medical experts in clinical text summarization. Nature Medicine. 2024;30(4):1134–42.

3. Tai-Seale M, Baxter SL, Vaida F, Walker A, Sitapati AM, Osborne C, Diaz J, Desai N, Webb S, Polston G, Helsten T. AI-generated draft replies integrated into health records and physicians’ electronic communication. JAMA Network Open. 2024;7(4):e246565..

4. Hager P, Jungmann F, Holland R, Bhagat K, Hubrecht I, Knauer M, Vielhauer J, Makowski M, Braren R, Kaissis G, Rueckert D. Evaluation and mitigation of the limitations of large language models in clinical decision-making. Nature medicine. 2024;30(9):2613–22.

5. Leung TI, Coristine AJ, Benis A. AI Scribes in Health Care: Balancing Transformative Potential With Responsible Integration. JMIR Medical Informatics. 2025;13(1):e80898.

6. Bracken A, Reilly C, Feeley A, Sheehan E, Merghani K, Feeley I. Artificial Intelligence (AI)–Powered Documentation Systems in Healthcare: A Systematic Review. Journal of Medical Systems. 2025;49(1):28. doi: 10.1007/s10916-025-02157-4.

7. Coiera E, Kocaballi B, Halamka J, Laranjo L. The digital scribe. NPJ Digit Med. 2018;1:58. doi: 10.1038/s41746-018-0066-9.

8. Balloch J, Sridharan S, Oldham G, Wray J, Gough P, Robinson R, Sebire NJ, Khalil S, Asgari E, Tan C, Taylor A. Use of an ambient artificial intelligence tool to improve quality of clinical documentation. Future Healthcare Journal. 2024;11(3):100157. doi: 10.1016/j.fhj.2024.100157.

9. Kernberg A, Gold JA, Mohan V. Using ChatGPT-4 to create structured medical notes from audio recordings of physician-patient encounters: comparative study. J Med Internet Res. 2024;26:e54419. doi: 10.2196/54419.

10. Biro J, Handley JL, Cobb NK, et al. Accuracy and safety of AI-enabled scribe technology: instrument validation study. J Med Internet Res. 2025;27:e64993. doi: 10.2196/64993.

11. Digital Scribes. Therapeutic Goods Administration, 14 August 2025. https://www.tga.gov.au/how-we-regulate/manufacturing/manufacture-medical-device/manufacture-specific-types-medical-devices/artificial-intelligence-ai-and-medical-device-software/digital-scribes

12. Luccioni AS, Pistilli G, Sefala R, Moorosi N. “Bridging the Gap: Integrating Ethics and Environmental Sustainability in AI Research and Practice.” arXiv preprint arXiv:2504.00797 (2025).

13. Liu X, Glocker B, McCradden MM, Ghassemi M, Denniston AK, Oakden-Rayner L. The medical algorithmic audit. The medical algorithmic audit. The Lancet Digital Health 2022;4(5): e384–e397. doi: 10.1016/S2589-7500(22)00003-6

14. McCradden MD, London AJ, Gichoya JW, Sendak M, Erdman L, Stedman I, Oakden-Rayner L, Akrout I, Anderson JA, Farmer LA, Greer R [et al]. CANAIRI: The collaboration for translational artificial intelligence trials in healthcare. Nature Medicine. 2025;31(1). doi.org/10.1038/s41591-024-03364-1.

15. Stakeholder engagement framework. Australian Government, Department of Health. https://www.health.gov.au/sites/default/files/stakeholder-engagement-framework.pdf&sa=D&source=docs&ust=1758520760993921&usg=AOvVaw2_0R3ZOgxRv3lXugQ9YryS

16. Aboriginal Health Impact Statement Policy. South Australian Government 27 October 2023.https://www.sahealth.sa.gov.au/wps/wcm/connect/public+content/sa+health+internet/about+us/governance/policy+governance/policies/aboriginal+health+impact+statement+policy

17. Aboriginal Health Plan 2024-2028. Women’s and Children’s Health Network. https://cdn.wchn.sa.gov.au/downloads/WCHN/aboriginal-health/Our-Aboriginal-Health-Plan-At-A-Glance-2024-2028.pdf&sa=D&source=docs&ust=1758520760994452&usg=AOvVaw1cOhcVB0AfzUX9U-HYVw-F

18. Zack T, Lehman E, Suzgun M, Rodriguez JA, Celi LA, Gichoya J, Jurafsky D, Szolovits P, Bates DW, Abdulnour RE, Butte AJ. Assessing the potential of GPT-4 to perpetuate racial and gender biases in health care: a model evaluation study. The Lancet Digital Health. 2024;6(1):e12–22. doi: 10.1016/S2589-7500(23)00225-X.

19. Abid, Abubakar, Maheen Farooqi, and James Zou. “Large language models associate Muslims with violence.” Nature Machine Intelligence 3, no. 6 (2021): 461–463.

20. Omiye JA, Lester JC, Spichak S, Rotemberg V, Daneshjou R. Large language models propagate race-based medicine. NPJ Digital Medicine. 2023;6(1):195. doi: 10.1038/s41746-023-00939-z.

21. Pfohl SR, Cole-Lewis H, Sayres R, Neal D, Asiedu M, Dieng A, Tomasev N, Rashid QM, Azizi S, Rostamzadeh N, McCoy LG. A toolbox for surfacing health equity harms and biases in large language models. Nature Medicine. 2024 Dec;30(12):3590–600. doi: 10.1038/s41591-024-03258-2.

22. Morey K, Franks C, Pearson O, Glover K, Brown A. Research according to whom? Developing a South Australian Aboriginal and Torres Strait Islander health research accord. First Nations Health and Wellbeing-The Lowitja Journal. 2023;1:100003.

